# Transcriptome- and proteome-wide Mendelian randomization to prioritize therapeutic targets for coronary heart disease

**DOI:** 10.1101/2024.06.27.24309406

**Authors:** Liam Gaziano, Elias Allara, Claudia Giambartolomei, David Stacey, Jing Hua Zhao, Hesam Dashti, Tao Jiang, Scott C. Ritchie, Brian R Ferolito, Danielle Rasooly, Gina M. Peloso, Emanuele Di Angelantonio, Eleanor Wheeler, Maik Pietzner, Themistocles L Assimes, Peter WF Wilson, Kelly Cho, Krishna G Aragam, Stephen Burgess, John Danesh, Claudia Langenberg, Juan Pablo Casas, J Michael Gaziano, Alexandre C Pereira, Adam S Butterworth

## Abstract

Despite widespread use of drugs targeting traditional cardiovascular risk factors such as lipids and blood pressure, a high burden of coronary heart disease (CHD) remains, hence novel therapeutics are needed for people who harbor residual risk. Using transcriptomic and proteomic data to instrument 15,527 genes or proteins, we conducted systematic *cis-*Mendelian randomization (MR) and conditional colocalization analyses with a genetic meta-analysis involving nearly 300,000 CHD cases. We identified 567 targets with putative causal relevance to CHD, of which 69 were not identified in previous genetic discovery or MR studies and were the sole causal signal in that genomic region. To aid translation of our findings, we annotated results with up-to-date information on drugs acting on these targets. Our results revealed opportunities for drug repurposing and development prioritization. For example, we provide evidence that cilostazol, a drug that targets *PDE3A* and is currently used for claudication, could be repurposed for prevention of CHD.

## Introduction

There is a need to develop novel therapeutics to prevent CHD, which remains a leading cause of death in adults across the globe^1^. Although several safe and effective drugs exist for primary prevention of CHD, many adults experience residual hypertension or dyslipidemia following standard therapies^2,3^, suggesting a need to identify new approaches to target these traditional cardiovascular risk factors. Furthermore, an increasing proportion of myocardial infarctions occur in patients without traditional cardiovascular risk factors^4^, highlighting the need to identify and modulate novel pathways implicated in coronary atherosclerosis and thrombosis.

Around 50% of drug development failures occur due to lack of efficacy^5^, suggesting that the chosen target is not causally related to the outcome. Mendelian randomization (MR)^6^ analyses exploit the random allocation of genetic alleles through the population and avoid the pitfalls of relying on pre-clinical models that may not translate to human biology. Indeed, drugs that target proteins with supportive evidence from human genetic studies have a higher chance of gaining regulatory approval^7^.

Recently, large-scale proteomic profiling has led to the creation of extensive catalogs of genetic variants that influence circulating protein levels^8-11^. Similarly, the GTEx consortium^12^ has measured transcriptome-wide RNA levels in nearly 50 tissues to identify associated genetic variants. In combination, these resources can provide tissue-specific instruments to evaluate the causal relevance of the human proteome and transcriptome on CHD. To achieve this goal, we leverage transcriptomic and proteomic summary statistics to perform Mendelian randomization and colocalization analyses between more than 15,000 genes or proteins and the risk of CHD, utilizing a *de novo* genome-wide association study (GWAS) meta-analysis involving nearly 300,000 cases from the Million Veteran Program^13^, CARDIoGRAMplusC4D^14^ and FinnGen^15^.

## Results

We took a similar approach as our previous transcriptomic and proteomic MR analyses^16^, with the overall study design for the current analysis shown in **Figure 1**. To minimize the potential for horizontal pleiotropy (i.e. the CHD-associated variants acting through a different gene/protein to the one being instrumented), we used locally-acting (*cis*), conditionally-independent, protein quantitative trait loci (*cis*-pQTLs) to instrument levels of 1,497 plasma proteins assayed using the SomaScan V4 platform in the Fenland study^17^ (**Supplementary Table 1**). We also selected conditionally-independent, *cis* gene expression QTLs (*cis*-eQTLs) from Version 8 of the Genotype-Tissue Expression (GTEx) resource^12^ as instruments for 15,412 protein-coding genes (**Supplementary Table 2**). With these instruments, we performed MR using a GWAS meta-analysis of CHD that we conducted using results from European ancestry participants of the Million Veteran Program (MVP)^18^, the latest CARDIoGRAMplusC4D Consortium GWAS^14^, and the FinnGen biobank^15^ involving 296,537 CHD cases among 1,593,766 participants (**Supplementary Table 3**). 15,527 genes or proteins had at least one instrumental variant in at least one tissue, hence we set a Bonferroni-adjusted significance threshold at *P=*3.22×10^-6^ (0.05/15,527). A full list of MR results can be found in **Supplementary Table 4**. To reduce the possibility that MR associations arise due to confounding by linkage disequilibrium (LD), we additionally performed a pairwise conditional colocalization^19^ analysis between pQTL or eQTL summary statistics and CHD summary statistics for all significant MR results. We annotated results (those that passed both Bonferroni-adjusted P-value for MR and colocalization [PPH4>0.80] thresholds) with information about drugs that act on these targets using the OpenTargets^20^ and DrugBank^21^ resources (**Supplementary Table 5-7**), and assessed novelty and potential for horizontal pleiotropy (**Supplementary Table 8**). For specific targets of interest, we performed phenome-wide association scans of instrumental variants and colocalization to identify putative mechanisms, safety signals and alternative indications (**Supplementary Table 9**). Lastly, we conducted enrichment analyses to assess whether the potential causal genes for CHD occur more frequently in certain biological pathways (**Supplementary Table 10**).

**Figure 1.**
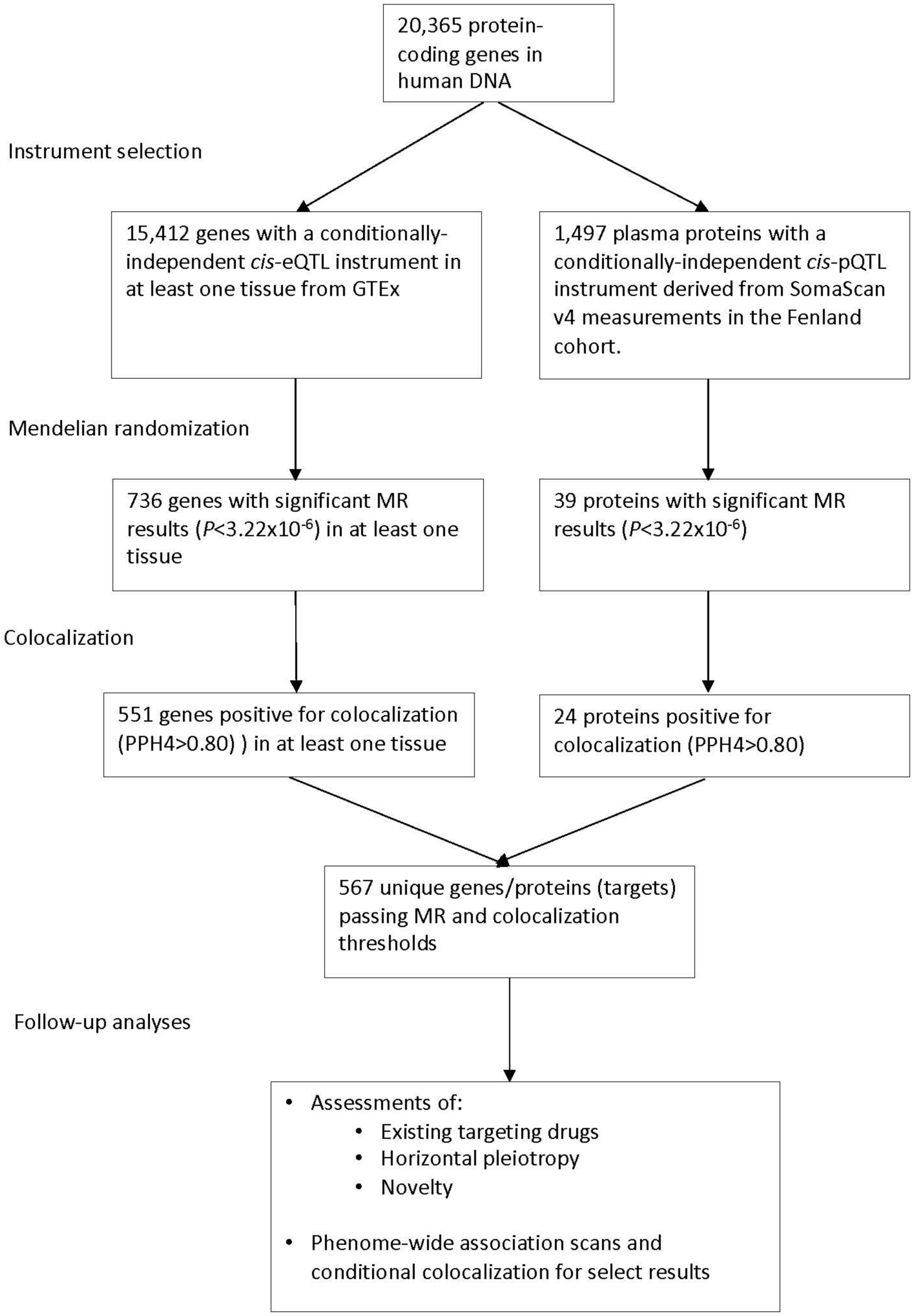
Overall study summary. We used *cis*-pQTLs to instrument 1,497 plasma proteins and *cis*-eQTLs to instrument 15,412 protein-coding genes to perform MR on a GWAS meta-analysis of CHD totaling 296,537 cases. For significant (*P<*3.22×10^-6^) MR results, we additionally performed colocalization using a pairwise conditional and colocalization (PWCoCo)^19^ approach between pQTL or eQTL summary statistics and CHD. Lastly, we annotated results (those that passed both Bonferroni-adjusted P-value for MR and colocalization [PP.H4>0.80] thresholds) according to whether an existing drug according status using the OpenTargets^37^ or DrugBank^21^.

Overall, we found 567 unique genes/proteins colocalizing with CHD. Using eQTL instruments, 736 genes passed the MR significance threshold (*P*<3.22×10^-6^) with 551 (74.9%) colocalizing (PPH4>0.8). Using pQTL instruments, 39 proteins passed MR significance with 24 colocalizing. Twenty of these were also tested using eQTL instruments, of which nine (*ABO*, *GSTT2B*, *IL6R*, *MST1*, *MXRA7*, *PCSK9*, *PDE5A*, *TMEM106A*, *TMEM106B*) also passed the colocalization threshold, and seven (*ABO*, *GSTT2B, MST1, MXRA7, PCSK9, PDE5A, TMEM106A*) had an eQTL signal in at least one tissue in the same direction as the plasma pQTL.

### Replication of past Mendelian randomization studies

We assessed whether we could replicate results from two transcriptome-/proteome-wide MR studies that have included CHD as an outcome, both of which used outcome summary statistics from the 2015 CARDIoGRAMplusC4D GWAS that included 60,801 CHD cases^22^. First, Zheng et al.^19^ used five^9,23-26^ publicly available pQTL resources to select instrumental variants, identifying *LPA* and *IL6R* with MR *P*<3.22×10^-6^ when restricting to results that used *cis*-instruments and colocalized. Here, we also identified *IL6R* (*P*=2.5×10^-22^, highest PPH4 in any tissue is 1.00) but could not assess *LPA*, as it is not measured on the SomaScan V4 panel. Second, Richardson *et al.*^27^ used *cis*-eQTL variants from GTEx Version 7 and eQTLGen^28^ as instruments and found 21 protein-coding genes with MR *P*<3.22×10^-6^ and *P*_HEIDI_ (a test for confounding by LD) >0.05, of which 20 were replicated here (*ABO, AIDA, ATP2B1, CARF, CELSR2, FAM117B, FES, GGCX, GUCY1A1, ICA1L, JCAD, LIPA, MIA3, MORF4L1, MRAS, NBEAL1, PHACTR1, PSRC1, SORT1, SWAP70*), suggesting that our framework identifies robust results. Only *VAMP8* from Richardson *et al.* did not colocalize in our analysis (highest PPH4 in any tissue was 0.33). In the *VAMP8* region, we instead identified *GGCX* (*P=*1.7x10^-52^, PPH4=1.00 in aortic artery tissue), which is targeted by the Anisindione, an anticoagulant occasionally used in those who cannot tolerate warfarin^29^. It is reassuring that our approach identified *GGXC* over *VAMP8* in that region, considering the biological role of *GGCX* in coagulation by activating vitamin K-dependent proteins and the relevance of coagulation to CHD.

### Assessment of horizontal pleiotropy

We considered an instrumental variant that influences expression of multiple genes / proteins as having the potential to introduce a form of horizontal pleiotropy that arises from shared regulation of nearby genes. Therefore, we grouped results with instrumental variants that are in close proximity (250kb) or are correlated (*r*^2^>0.2 in 1000 Genomes EUR). The 567 genes/proteins, hereafter referred to as “targets”, that passed MR and colocalization thresholds lie within 283 genomic regions. We allocated results into tiers based on the number of targets within a genomic region: Tier 1 includes 165 targets that are the sole result within a region to pass MR and colocalization thresholds, while the remaining 404 results (located within 116 regions) were annotated as Tier 2 (**Supplementary Table 8**). Tier 1 genes included several well-known to relate to CHD risk, such as *APOB, IL6R, IL1RN, LIPA*, and others previously identified by GWAS as the most likely causal gene in the region, such as *ITGA1, MYO9B,* and *SERPINA1^14^*. Because we tested all protein-coding genes in GTEx, Tier 1 results that are the only significant signal in the region are less likely to exhibit this form of horizontal pleiotropy. Conversely, since Tier 2 results contain multiple targets, it can be difficult to disentangle the gene responsible for driving the signal(s) in each region using MR and colocalization alone. For example, in one region on chromosome 1 (region #13 in **Supplementary Table 8**), there are four targets (*SARS1*, *CELSR2*, *PSRC1* and *SORT1*) that pass MR and colocalization thresholds. However, previous functional work has shown that *SORT1* is likely driving the signal in the 1p13.3 locus^30^, meaning results for *SARS1, CELSR2,* and *PSRC1* likely arose due to horizontal pleiotropy (**Supplementary** Figure 1).

### Novel putative therapeutic targets

We classified results as novel if they (1) were based on instrumental variants that were not correlated (*r*^2^<0.2 in 1000 Genomes EUR) or in close proximity (±250kb) with genome-wide significant (*P*<5×10^-8^) variants for CHD reported in Tcheandjieu *et al*.^18^, Aragam *et al*.^14^, or Supplementary Table 2 of Aragam *et al*.^14^ which includes 216 CHD-associated variants previously reported in the literature and (2) were not reported in Zheng *et al*.^19^ or Richardson *et al*.^27^. We classified 184 targets as novel, 69 of which were classified also as Tier 1 (**Figure 2**, **Supplementary Table 8**).

**Figure 2.**
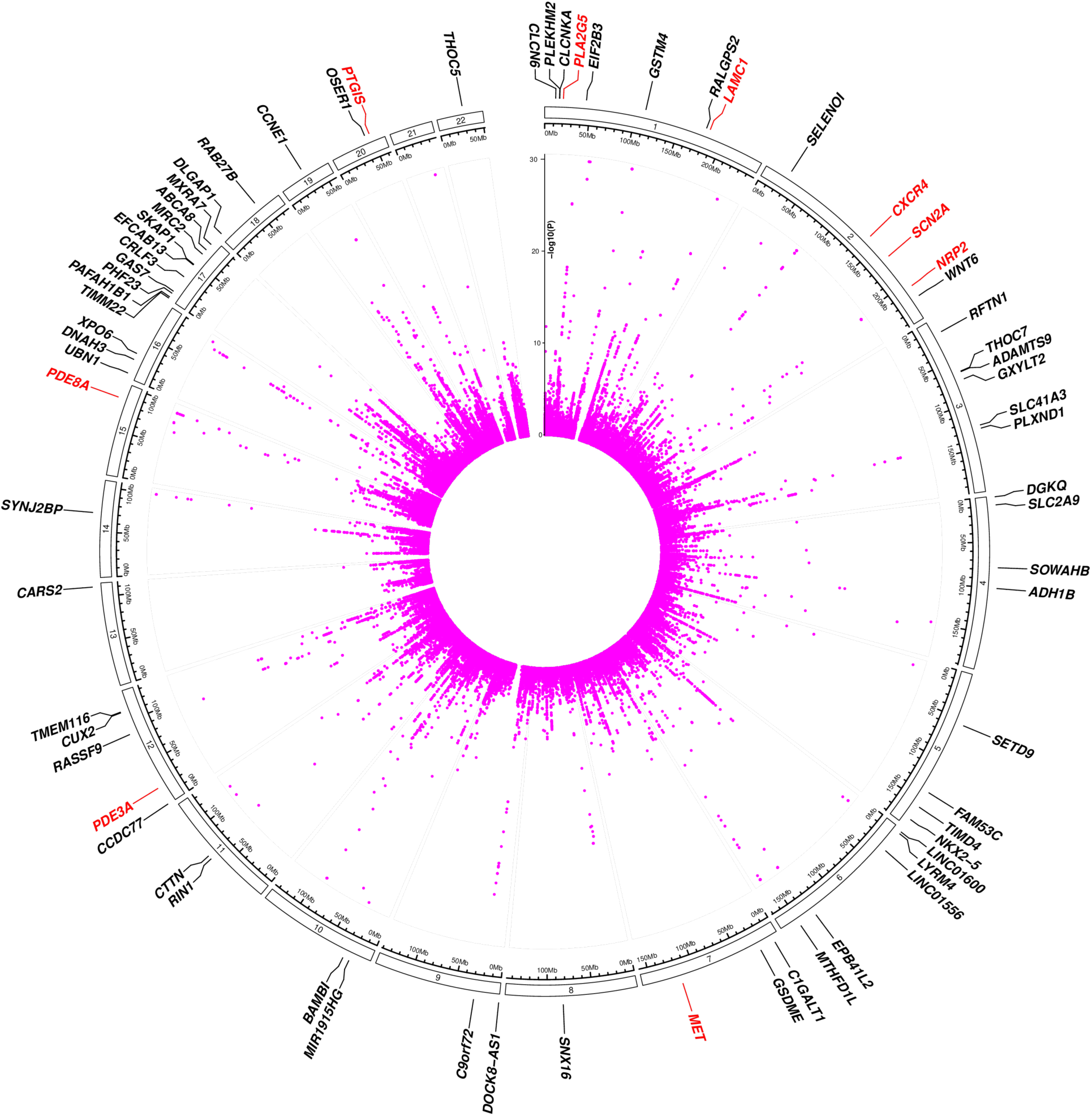
Circos plot from Mendelian randomization and colocalization analysis. Targets that are labeled are Tier 1 and novel results that passed MR (*P*<3.22×10^-6^) and colocalization thresholds (PPH4>0.8). Tier 1 results are defined as those that are not in close proximity (+250kb) or correlated (*r*^2^>0.2 in 1000 Genomes EUR) with any other result that passed MR and colocalization thresholds. Results were defined as novel if they (1) were based on instrumental variants that were not correlated (*r*^2^<0.2 in 1000 Genomes EUR) or in close proximity (±250kb) with genome-wide significant (*P*<5×10^-8^) variants for CHD reported in Aragam *et al*.^14^ or Tcheandjieu *et al*.^18^ or Supplementary Table 2 of Aragam *et al*.^14^ which includes CHD-associated variants previously reported in the literature and (2) were not previously reported in Zheng *et al*.^19^ or Richardson *et al*.^27^ Targets with existing drug according to either OpenTargets^37^ or DrugBank^21^ are labeled in red, targets without a current drug are labeled in black.

### Targets with existing drugs

Of the 567 targets that passed MR and colocalization thresholds, 30 had drugs developed that act on these targets according to DrugBank^21^ (**Supplementary Table 5**) and 32 according to OpenTargets^20^ (**Supplementary Table 6**), of which 19 from both. Forty-three targets had an existing drug according to either DrugBank or OpenTargets and the remaining 524 targets were classified as not currently having a drug (**Supplementary Table 7**). Of the 43 targets with existing drugs, several are involved in well-known vascular pathways such as lipid metabolism (e.g., *PCSK9, LPL, APOC3, NPC1L1, CETP, ABCA1*), hemostasis (e.g., *F11, FN1, PDE3A, PIK3CB, TFPI, KLKB1, PDE8A, GGCX*) and inflammation (e.g. *IL6R, IL23A, MIF*). Some of these - such as *PCSK9*^31^*, NP1CL1*^32^, and *CETP*^33^ - have already been shown to be effective therapeutic targets for CHD in clinical trials, while others like *IL6R* are currently being tested^34^.

#### Repurposing

Our results also reveal notable repurposing opportunities. An example is the novel, Tier 1 result for PDE3A, which is targeted by several inhibitory drugs for a wide range of indications, including thrombotic events and respiratory diseases (**Supplementary Table 5 and 6**). Higher genetically-predicted expression of *PDE3A* was positively associated with CHD (*P*=2.25×10^-7^) using a colocalizing (PPH4=0.99) eQTL instrumental variant (rs7488772) in subcutaneous adipose tissue, suggesting that therapeutic inhibition might be beneficial. A phenome-wide association scan with colocalization (PheWAS-coloc) for rs7488772 showed that higher genetically-predicted expression of *PDE3A* was associated with higher levels of several cardiometabolic risk factors, including systolic blood pressure, triglycerides, and glycated hemoglobin (**Figure 3** and **Supplementary Table 9**).

**Figure 3.**
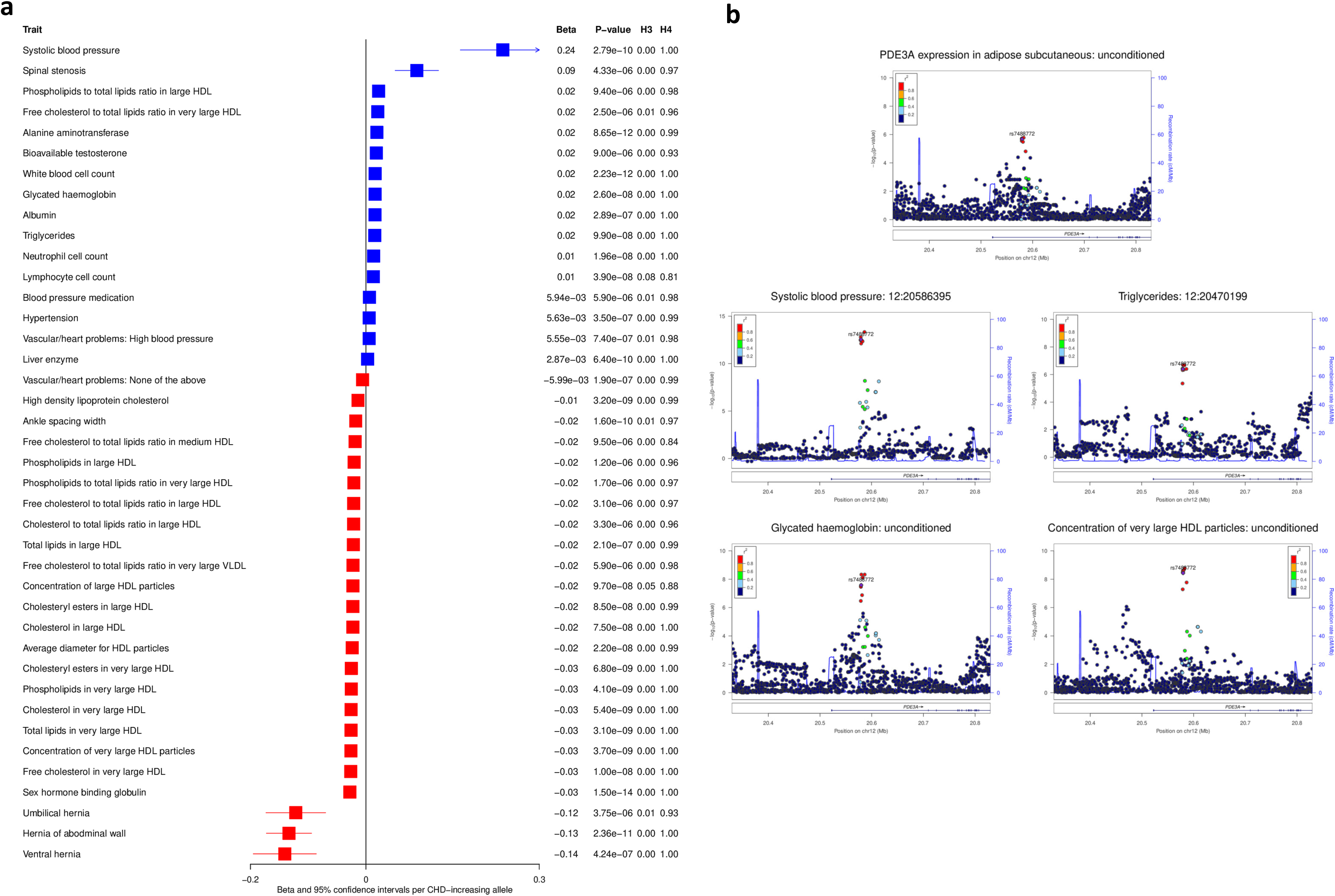
Phenome-wide scan and colocalization results for *PDE3A*. Results are shown for scans in MR Base^66^ of rs7488772, the instrumental variant for *PDE3A* in adipose subcutaneous tissue. **(a)** results for all genetic associations with *P*<1×10^-5^ (aligned by CHD-increasing allele; positive betas are shown in blue and negative betas in red; if the same trait is estimated in more than one dataset, this plot shows the one with the smallest p-value) and conditional colocalization PPH4≥0.8 (if a trait colocalizes with more than one variant after conditional analysis, this plot shows the highest PPH4), and **(b)** LocusZoom^67^ plots for selected cardiometabolic traits. Results from colocalization analyses using marginal summary statistics are labeled as ‘unconditioned’. Results from conditional colocalization show the conditioning variant in the plot title.

#### Identifying safety concerns for non-cardiovascular drugs

We also found an inverse association between higher genetically-predicted expression of *CXCR4* (instrumented by rs10171574) and risk of CHD (*P*=9.52x10^-7^) (**Supplementary Table 5**). PheWAS-coloc showed associations of the CHD-increasing (and *CXCR4*-decreasing) allele with higher neutrophil percentage of white blood cells and lower lymphocyte percentage of white blood cells (**Supplementary** Figure 2 and **Supplementary Table 9**). These associations are consistent with the role of *CXCR4* inhibitors such as plerixafor, which is commonly used in stem cell transplantation to mobilize granulocytes and their precursors into the blood stream^35^. Our findings suggest that *CXCR4* inhibitors (e.g. plerixafor and uloplucumab) may increase risk of CHD in an on-target fashion, raising potential safety concerns for long-term use.

#### Disentangling effects for drugs with multiple targets

One therapeutic target, *PLA2G5*, which encodes secretory phospholipase A2 group V (sPLA2-V), is inhibited by Varespladib methyl. We found that higher genetically-predicted *PLA2G5* expression, a Tier 1 result, was associated with higher risk of CHD (*P*=3.18×10^-6^) (**Figure 4**) using an instrumental variant (rs12408798) in aortic artery tissue (PPH4=0.99). However, Varespladib methyl did not reduce coronary events in ∼5000 patients with acute coronary syndrome in the VISTA-16 trial^36^, and even showed an increased risk of myocardial infarction. Varespladib methyl also targets isoenzymes sPLA2-IIA (encoded by *PLA2G2A*) and sPLA2-X (encoded by *PLA2G10*)^37^. Here, eQTL instruments for *PLA2G2A* expression appeared in multiple tissues, none of which were associated with CHD (*P*>0.52), while there was no eQTL instrument for *PLA2G10* in any tissue nor a pQTL instrument.

**Figure 4.**
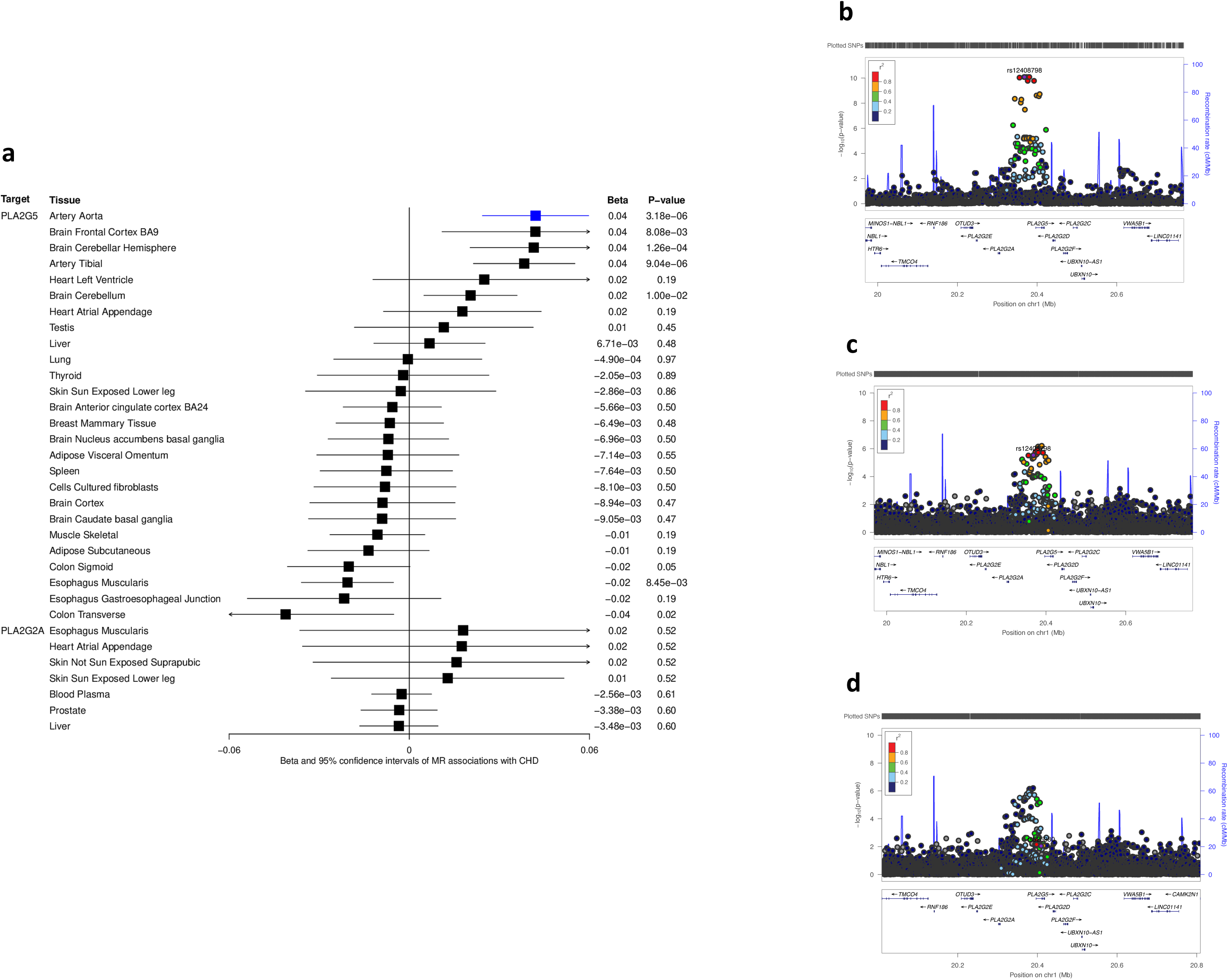
Results for *PLA2G5* and *PLA2G2A*. **(a)** Mendelian randomization results for *PLA2G5* and *PLA2G2A* in all tissues where an instrument exists. Only results for *PLA2G5* in aortic artery tissue (highlighted in blue) passed the P-value significance threshold and was tested for colocalization. All results are from eQTL except for *PLA2F2A* / blood plasma which are derived from pQTL. Regional association statistics for **(b)** gene expression of *PLA2G5* in aortic artery tissue from GTEx and **(c)** coronary heart disease. **(a)** and **(b)** show the pairwise correlation (1000G European ancestry) for each variant with rs12408798, the instrumental variant used in the present study. **(a)** and **(b)** colocalize at PPH4=0.98. **(d)** Local association plot of the *PLA2G5* region for coronary heart disease, showing the correlation for rs525380, the instrumental variant for *PLA2G5* expression used in the previous study Holmes *et al*.^53^ The association for rs525380 and CHD was *P*=0.20 in Holmes *et al*.^53^ and *P*= 0.009 in the present study.

### Targets currently without a drug

The 524 targets for which drugs do not currently exist can help prioritize drug development programs. For example, we found positive associations for genetically-predicted *ABCA8* expression with risk of CHD (*P*=6.50×10^-7^), suggesting ATP binding cassette subfamily A member 8 as a potential therapeutic target. PheWAS-coloc for *ABCA8* (instrumented by rs34931250 in several tissues) showed positive associations with LDL-cholesterol concentration and Apolipoprotein B, and inverse associations with HDL-cholesterol and Apolipoprotein-A1 (**Figure 5** and **Supplementary Table 9**), suggesting pro-atherogenic lipids and poor cholesterol efflux as the potential mediators.

**Figure 5.**
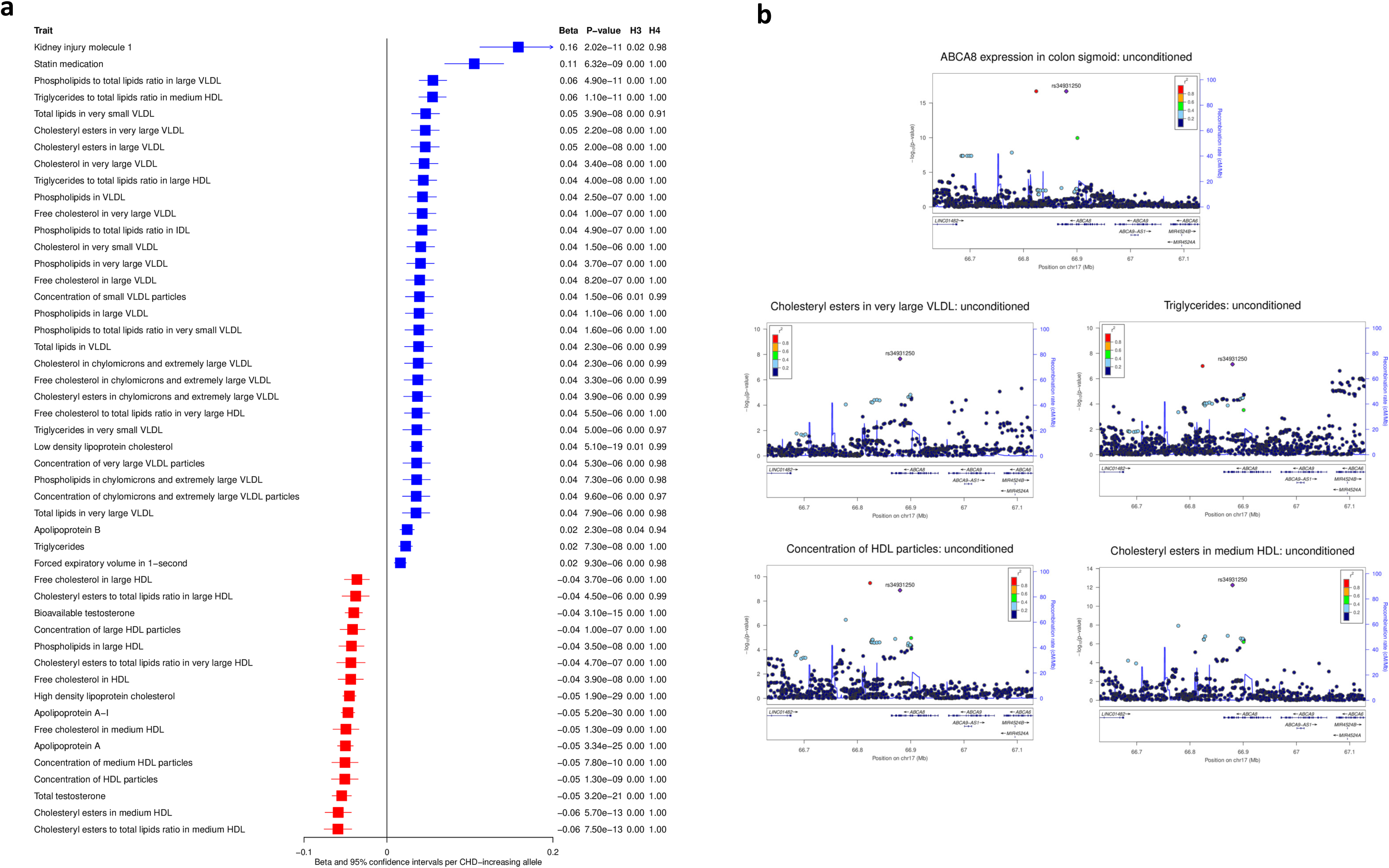
Phenome-wide scan and colocalization results for *ABCA8*. Results are shown for scans in MR Base^66^ of rs34931250, the instrumental variant for *ABCA8*. **(a)** results for all genetic associations with *P*<1×10^-5^ (aligned by CHD-increasing allele; positive betas are shown in blue and negative betas in red; if the same trait is estimated in more than one datasets, this plot shows the one with the smallest p-value) and conditional colocalization PPH4≥0.8 (if a trait colocalizes with more than one variant after conditional analysis, this plot shows the highest PPH4), and **(b)** LocusZoom plots for selected cardiovascular traits. Results from colocalization analyses using marginal summary statistics are labeled as ‘unconditioned’.

### Pathway and gene ontology term enrichment analysis

To determine whether our set of 567 putative causal genes for CAD was over-represented in specific biological mechanisms or pathways, we performed enrichment analyses using Enrichr^38-40^. Overall, we found significant (*P_adj_*<0.05) evidence of enrichment for 25 Kyoto Encyclopedia of Genes and Genomes (KEGG) pathways or gene ontology (GO) terms, with the highest significance observed for lipid-related categories (**Supplementary Table 10**). This included the KEGG pathways “cholesterol metabolism” (*P*=8.85x10^-10^, odds ratio [OR]=12.32) and “fat digestion and absorption” (*P*=2.37×10^-5^, OR=7.95), as well as the GO terms “lipoprotein particle remodeling” (*P*=3.35×10^-5^, OR=12.24) and “sterol transfer activity” (*P*=1.49x10^-4^, OR=12.36). Furthermore, Enrichr also highlighted (*P_adj_*<0.05) GO terms relating to endothelial cell migration (*P*=2.71×10^-5^, OR=5.19), focal adhesion (*P*=3.00x10^-4^, OR=2.33), and cell-substrate junctions (*P*=4.03x10^-4^, OR=2.28). Thus, these analyses revealed a highly significant enrichment of genes involved in CAD-related biological pathways and mechanisms.

We then performed a separate enrichment analysis focusing specifically on our set of 165 Tier 1 putative causal genes. These analyses highlighted 51 significant (*P_adj_*<0.05) KEGG pathways or GO terms (**Supplementary Table 11**), double the number observed in the previous enrichment analysis of all 567 candidates and which again included the KEGG pathways “cholesterol metabolism” (*P*=3.39×10^-6^, OR=16.97) and “fat digestion and absorption” (*P*=4.29×10^-4^, OR=12.61). The Tier 1 analysis also highlighted a major theme related to cell migration, motility, and chemotaxis, while reiterating an enrichment of gene products localized to focal adhesions (*P*=4.18x10^-6^, OR=4.84) and cell-substrate junctions (*P*=5.29x10^-6^, OR=4.73). Together, this analysis, confined to the 165 Tier 1 genes, highlights pathways that influence CAD risk by regulating spatial arrangements and interactions between cells at the arterial wall, which is consistent with current models of atherosclerosis^41^.

## Discussion

Using instruments derived from gene expression in multiple tissues and plasma protein levels, we identified 567 targets within 283 genomic regions with a putative causal relevance to CHD. We annotated the findings with information on existence of current targeting drugs to aid translation of our findings into actionable interventions and followed-up specific results with phenome-wide scans and colocalization to identify possible mechanisms or on-target safety concerns. While previous studies have used transcriptomic and proteomic genetic instruments on CHD^19,27^, none have been applied to a GWAS of this size. As a result, this analysis increased the number of genes/proteins with putative causal relevance to the development of CHD, with 69 novel, Tier 1, eQTL-CHD or pQTL-CHD pairs.

These results can help establish novel therapeutics for prevention of CHD in several ways. First, we identified possible opportunities for drugs currently on the market, that might be repurposed. PDE3A is targeted by multiple small molecules, two of which (milrinone and amrinone) were tested in overt heart failure but displayed null or harmful effects^42-44^, and one, cilostazol, which is currently used for intermittent claudication. Cilostazol has antiplatelet and vasodilatory effects^45^ by inhibiting PDE3A’s catalysis of inactive adenosine monophosphate (AMP) from cyclic (cAMP). We found higher genetically-predicted *PDE3A* expression to be positively associated with CHD and a number of cardiovascular risk factors. Some of the PheWAS-coloc results for the *PDE3A* instrumental variant have been additionally corroborated by human trials of cilostazol. For example, trials have found that cilostazol reduces triglycerides and increases HDL-cholesterol^46,47^. Cilostazol, in addition to its antiplatelet effects, also acts as a vasodilator, so the PheWAS-coloc result for systolic blood pressure is perhaps not surprising, especially considering cilostazol has been shown in trials to reduce ankle brachial index^48^. The congruence between existing trial data and the PheWAS-coloc analysis provides evidence that the instrumental variant for *PDE3A* expression is accurately mimicking the on-target effects of cilostazol. Our findings additionally corroborate previous findings for PDE3B (a protein with similar function as PDE3A and is also targeted by Cilostazol^49^), which found loss of function mutations in *PDE3B* are associated with reductions in CHD, decreased triglycerides, and increased HDL-C^50^.

Cilostazol has also been shown to reduce secondary ischemic stroke by 32% in a meta-analysis of trials^51^. It is often tested against antiplatelet drugs, like aspirin, clopidogrel, or a combination of the two, and has shown superiority in preventing bleeding events as well, including a 57% reduction in hemorrhagic stroke^51^. While aspirin and clopidogrel reduce CHD in primary prevention, there is debate about whether the increased bleeding risk outweighs the benefits^52^. Therefore, large trials of cilostazol for primary prevention of CHD may be warranted to determine if it provides a safer and more effective antiplatelet strategy than aspirin or clopidogrel.

Second, our findings can help clear up questions surrounding drugs that target multiple proteins. Varespladib inhibits isoenzymes sPLA2-V (encoded by *PLA2G5*), sPLA2-IIA (encoded by *PLA2G2A*) and sPLA2-X (encoded by *PLA2G10*). All three of these genes have been assessed previously in MR studies for CHD and all have been null^53-55^. The previous study instrumented gene expression of *PLA2G5* with rs525380 in aorta adventitia tissue from 133 participants of the Advanced Study of Aortic Pathology (ASAP). ASAP participants were genotyped using the Illumina Human 610 W-Quad Bead array without imputation. The variant from Holmes *et al*., rs525380, is in weak LD (*r*^2^=0.28 in 1000 Genomes European ancestry) with the one used here for *PLA2G5*, and this may explain the different results (**Figure 4**). We replicated a separate study that found no association between genetically-predicted *PLA2G2A* and CHD^54^ that used an identical instrumental variant as the present analysis (rs11573156). A previous MR analysis on sPLA2-X suffered from weak instruments for *PLA2G10* expression^55^ (similarly, there were no significant eQTLs for *PLA2G10* in GTEx), and without strong genetic instruments it is difficult to understand its causal relevance to CHD through MR.

The phase 3 VISTA-16 trial found that Varespladib had no effect on a composite outcome of cardiovascular mortality, nonfatal MI, nonfatal stroke, or unstable angina in those with acute coronary syndrome^36^ but did find an increase in myocardial infarction. This may be explained by Varespladib’s inhibition of sPLA2-X, which showed antiatherogenic effects in mouse models^56^. Indeed, Varespladib is five times more active for sPLA2-X than sPLA2-V, with half maximal inhibitory concentrations of 15 nM for sPLA2-X versus 77 nM for sPLA2-V^57^. In aggregate, these results suggest that specific inhibition of sPLA2-V without inhibition of sPLA2-IIA or sPLA2-X could be an effective intervention for CHD prevention.

Third, MR results for non-druggable targets can help to indicate which targets should be carried forward for development. We found that higher levels of *ABCA8* gene expression decrease HDL-cholesterol levels and increase LDL-cholesterol levels and risk of CHD, suggesting that modulating ABCA8 could prevent CHD, potentially through either reductions in Apo-B-containing lipoproteins or improved cholesterol efflux. Our findings for HDL-cholesterol are discordant with studies of deleterious *ABCA8* mutations and *ABCA8* knock-out mice, which have shown *decreased* HDL-cholesterol levels ^58-60^. Combined with the biological role of ABCA8 and other ATP binding cassette proteins^58^, and the congruence between the effect of the instrumental variant on ApoB-containing lipoproteins and CHD, these findings suggest that it is actually reduced ABCA8 protein levels that increase ApoB-containing lipoproteins and risk of CHD. This highlights one of the challenges of transcriptomic-derived instruments, in that there are several reasons why expression of a gene may be elevated. For example, gene expression may be upregulated as a compensatory mechanism for a variant that causes decreased function of a protein through changes in key amino acids, splicing, or other mechanisms. It is possible such a phenomenon is occurring for *ABCA8*, particularly because the instrumental variant lies within a splice region^61^. More in-depth evaluation of ABCA8 and the many other potentially causal targets instrumented by gene expression are warranted to determine the desired direction of therapeutic modulation.

Lastly, our findings can help identify on-target effects of drugs that may increase the risk of CHD, suggesting potential safety concerns We found an association between greater expression of *CXCR4* and lower risk of CHD, which is consistent with a previous study in hyperlipidemic mice^62^. Additionally, CXCL12, the natural ligand for CXCR4, shows a positive, Tier 1, association for CHD in our analysis. *CXCR4*, which encodes C-X-C chemokine receptor type 4, is the target of multiple marketed medications, including Plerixafor and Ulocuplumab, and our findings suggest that these drugs could increase the risk of CHD in an on-target fashion. CXCR4 inhibitors are currently used in stem cell transplant donors to elicit more stem cells into the peripheral blood and this transient use is unlikely to have a lasting harmful impact on CHD risk. However, they have been considered for more long-term use for other conditions^63-65^, and our findings recommend evaluation of cardiovascular safety in such settings. Other drug development programs can similarly use our findings to better predict whether a drug for any disease could have a harmful on-target effect on CHD.

The major limitation of our analytical approach is that within 116 loci there were multiple targets that passed MR and colocalization thresholds. Tier 2 results that lie within the same genomic region, like the *SORT1*, *CELSR2* and *PSRC1* locus, provide evidence that even *cis*-instruments may exhibit horizontal pleiotropy, perhaps through shared regulation of multiple local genes or linkage disequilibrium, making it difficult to determine the true causal target(s) in a region. Although the approach taken here equally prioritizes these genes, functional work has suggested that *SORT1* is likely driving the signal in the 1p13.3 locus^30^. Conversely, though, due to our comprehensive testing of all protein-coding genes in GTEx, Tier 1 results that are the only result in the region are more likely to be the causal gene driving the association.

There are other limitations as well. This study was undertaken in almost exclusively European ancestry, so other studies in diverse ancestries are warranted. The pQTL MR only covers a small proportion of the human proteome and instrumenting plasma protein levels may not always provide the most relevant tissue in the context of CHD. The eQTL MR analysis used instruments for gene expression which may not translate to effects on protein levels in a relevant cell type. Lastly it can be difficult through MR, especially with gene expression data, to determine how much inhibition or activation of a certain protein is required for a meaningful reduction in CHD.

Our study has several strengths. We conducted a *de novo* GWAS meta-analysis involving nearly 300,000 CHD cases, providing a well-powered outcome dataset. We performed conditional colocalization analysis to robustly test for pleiotropy, avoiding the one-signal-per-locus limitation of the traditional colocalization approaches. Annotations of results with up-to-date information on the druggable genome help facilitate translation into actionable interventions. Further investigations with phenome-wide scans with colocalization on instrumental variants can identify possible on-target safety concerns as well as elucidate potential mechanisms for CHD pathogenesis.

In summary, our transcriptomics and proteomics analysis identified 69 novel, Tier 1 targets with putative causal relevance for CHD. We show opportunities for drug repurposing and development prioritization. Additionally, our results can be used for understanding CHD safety of current and future drugs indicated for non-cardiovascular diseases.

## Author contribution

L.G., J.P.C., A.C.P., and A.S.B. conceptualized the study. E.W. M.P. and C.L. generated pQTL information. S.C.R., T.J., K.A., T.J.A., P.W.F.W., K.C. assisted in preparation of CHD summary statistics. L.G. performed the Mendelian randomization analysis. C.G. performed conditional colocalization analysis on CHD. E.A. performed phenome-wide scans of instrumental variants with conditional colocalization. D.S. performed pathway enrichment analysis. L.G., J.H.Z., and E.A. generated figures. L.G., E.A., A.C.P., and A.S.B. wrote the manuscript. A.S.B. oversaw all analyses. All authors contributed to study design and provided editorial comments.

## Methods

### Selecting genetic instruments

First, we selected conditionally-independent *cis*-pQTLs for plasma proteins reported in Pietzner *et al*. (www.omicscience.org)^8^ Briefly, protein levels were measured by SomaLogic Inc (Boulder, Colorado, US) with the SomaScan® V4 platform in 10,708 European-ancestry individuals from the Fenland cohort^17,68^. This technique uses DNA-aptamers called SOMAmers, oligonucleotide strands that fold and bind to proteins with high specificity, and association analyses were performed for 5,210 SOMAmers. We removed pQTLs with minor allele frequency (MAF)<0.01, *trans*-pQTLs and SOMAmers that measured more than one protein.

For eQTL instruments, we downloaded a file provided by the Genotype-Tissue Expression (GTEx) Consortium version 8^12^ (https://storage.googleapis.com/gtex_analysis_v8/single_tissue_qtl_data/GTEx_Analysis_v8_eQ TL_independent.tar) that included all conditionally independent *cis*-eQTLs for expression of all genes in 49 tissues. We excluded eQTLs for non-protein-coding genes (i.e. those that do not have a UniProt ID), *P* > 5×10^-8^ and MAF<0.01. We also removed eQTLs, pQTLs or genes that lie within the human leukocyte antigen (HLA) region (chr6:29691116-33054976), due to the complicated LD structure in that region.

### Generation of outcome summary statistics

To generate outcome summary statistics for CHD, we meta-analyzed results from three non-overlapping sources: The Million Veteran Program (MVP)^18^, CARDIoGRAMplusC4D Consortium^14^, and the FinnGen biobank^15^. MVP is an ongoing biobank recruiting from 63 Veterans Affairs (VA) facilities across the United States^13^. In MVP, we utilized a GWAS that consisted of European ancestry individuals. The CARDIoGRAMplusC4D Consortium is a GWAS meta-analysis on CHD from many contributing studies, primarily of European ancestry^14^. FinnGen is a consortium of biobanks within Finland that includes participants linked to electronic health data. For FinnGen, we downloaded summary statistics for the I9_ISCHHEART outcome (gs://finngen-public-data-r3/summary_stats/finngen_r3_I9_ISCHHEART.gz) from release 3, which defined CHD as ICD-10 I20-I25. We meta-analyzed summary statistics from the three data sources using METAL software^69^ with fixed-effects and inverse-variance weighting.

### Mendelian Randomization

We used the TwoSampleMR R package (https://github.com/MRCIEU/TwoSampleMR) to estimate MR effects between instruments and CHD. Inverse variance-weighted MR with fixed-effects was used for instruments with more than one variant, and the Wald ratio method for instruments with one variant. For instruments with more than one variant, we additionally assessed heterogeneity across the variant-level MR estimates and reported the Cochran Q *P* value.

### Conditional Colocalization

Significant MR associations can arise from a strong association between an instrumental variant and an outcome that is not causal, but in LD with a peak variant (i.e. an MR estimate can be due to confounding by LD and not due to a shared causal variant). To address this, we performed colocalization between the pQTL or eQTL summary statistics and CHD for all results that passed the MR *P*-value threshold. Additionally, when performing colocalization, false negatives can arise from interference of a strong independent signal, because conventional colocalization methods traditionally assume one causal variant within the tested region. To address this, we performed pairwise conditional colocalization^19^, which requires conditional summary statistics for each region. Conditional analyses for pQTL data from Pietzner *et al*.^8^ were conducted using GCTA-COJO^70^ and a reference panel of genotypes from the Fenland cohort. GCTA-COJO is a stepwise model selection procedure to select independently associated SNPs using the LD structure to account for the correlation between SNPs. For eQTLs, we generated summary statistics using raw genotype data from European ancestry participants following the procedure outlined by the GTEx Consortium. Briefly, after filtering the genotypes (MAF<0.01, HWE<0.000001, and no ambiguous SNPs), GWAS between variants and gene expression was performed adjusting for the top 5 principal components, PEER factors, sequencing platform and sex. Conditional analysis for gene expression were performed with raw genotype data by iteratively adjusting for the most strongly associated variant in a region. Conditional analysis for CHD summary statistics were performed using GCTA-COJO with a 1000G^71^ European ancestry reference panel.

Pairwise colocalization was performed using the COLOC R package^72^ on regions ±200kb around the instrumental variant with at least 50 SNPs matching across the two datasets. The prior probability of each SNP that is causal to only one of the traits (PPH3) was set to 1 × 10^−5^ and causal to both traits (PPH4) was set to 1 × 10^−6^. A posterior probability of a shared causal variant (PPH4) of ≥80% for any of the conditional iterations was considered strong evidence of colocalization.

### Drugs against identified targets

Targets that passed MR and colocalization thresholds were annotated with information on drugs against targets from by OpenTargets^20^ and DrugBank^21^. We used DrugBank release 5.1.10, (https://go.drugbank.com/releases) and restricted to known pharmacologically active drug-target pairs.

### Assessment of horizontal pleiotropy

Valid instrumental variants for MR must meet the exclusion restriction requirement, which assumes that the variant only affects the outcome through the instrumented exposure. A variant is an invalid instrument if it alters expression of other genes or proteins that affect the outcome through distinct biological/causal pathways (horizontal pleiotropy). In the case that results for multiple genes/proteins in a region colocalize with the same CHD signal, it can be difficult to disentangle which is truly influencing CHD risk. Therefore, we considered a variant that influences expression of nearby genes through shared regulation as possibly introducing horizontal pleiotropy and grouped results into regions based on instrumental variants that are in close proximity (+250kb) or correlated (*r*^2^>0.2 in 1000 Genomes EUR). Because we tested all protein-coding genes in GTEx, results that are the only significant signal in a region are less likely to exhibit horizontal pleiotropy. Therefore, we allocated results into tiers based on the number of results within a genomic region, where Tier 1 results are the sole result within a region to pass MR and colocalization thresholds, and Tier 2 results contain multiple signals within a region.

### Phenome-wide scans of instrumental variants

For some results that passed both MR and colocalization thresholds, we performed Phenome-wide association scan with colocalization (PheWAS) of instrumental variants. Associations between instrumental variants with disease and traits can mimic on-target effects of therapeutic targeting observed in trials. This can provide confidence that instrumental variants are correctly mimicking on-target effects of a drug. Additionally, a PheWAS analysis can identify possible harmful effects of protein modulation or provide mechanistic insights. Therefore, we queried instrumental variants for associations with diseases and traits in the MRbase PheWAS platform^66^, a database that allows for PheWAS across a wide range of publicly available datasets. For PheWAS associations with *P* <1×10^−5^ we also performed pairwise conditional colocalization between eQTL and traits regional summary stats, to help ensure those associations are not due to confounding by LD. For this, we utilized the PwCoCo v. 1.0 (https://github.com/jwr-git/pwcoco/tree/pwcoco-v1.0) tool^73^, using LD information from the UK Biobank after restricting the analysis sample to 367,641 unrelated European-ancestry participants. We set the prior probabilities that each SNP was causal to either of the traits (PPH3) and causal to both traits (PPH4) to 1 × 10^−5^ and 1 × 10^−6^, respectively. We considered a posterior probability of a shared causal variant (PPH4) of ≥80% as strong evidence of colocalization. For inspection with LocusZoom^67^ (http://genome.sph.umich.edu/wiki/LocusZoom_Standalone), we also exported conditioned genetic associations using the "--out-cond" option in PwCoCo.

### Pathway and gene ontology term enrichment analysis

Pathway and functional term enrichment analyses were performed with Enrichr (https://maayanlab.cloud/Enrichr/)^38-40^ by inputting the HGNC symbols for all putative causal targets as a single gene set. Briefly, Enrichr uses the Fisher’s exact test to assess whether the putative causal gene set was enriched with genes assigned to a given GO term or KEGG pathway relative to a permuted background gene set. An odds ratio significantly greater than one is indicative of an enriched term or pathway. Correction for multiple testing (*P_adj_*) was conducted using the Benjamini-Hochberg procedure.

## Data Availability

All data produced in the present work are contained in the manuscript or supplementary files.

## Acknowledgements

Scott C. Ritchie was funded by a British Heart Foundation (BHF) Programme Grant (RG/18/13/33946) and by the National Institute for Health and Care Research (NIHR) Cambridge BRC (BRC-1215-20014; NIHR203312) [*] * The views expressed are those of the authors and not necessarily those of the NIHR or the Department of Health and Social Care. Emanuele Di Angelantonio holds an NIHR Senior Investigator Award. Stephen Burgess is supported by the Wellcome Trust (225790/Z/22/Z) and the United Kingdom Research and Innovation Medical Research Council (MC_UU_00002/7). John Danesh holds a British Heart Foundation Professorship and an NIHR Senior Investigator Award. Adam Butterworth and Claudia Langenberg were supported by BHF-DZHK grant funding “Prot4CVD: Translational proteomics for cardiovascular diseases: from population prediction to clinical and therapeutic applications” (FKZ 81X2100281). The BHF Cardiovascular Epidemiology Unit was supported by core funding from the NIHR Blood and Transplant Research Unit in Donor Health and Genomics (NIHR BTRU-2014-10024), the NIHR BTRU in Donor Health and Behaviour (NIHR203337), the UK Medical Research Council (MR/L003120/1), the British Heart Foundation (SP/09/002; RG/13/13/30194; RG/18/13/33946) and NIHR Cambridge BRC (BRC-1215-20014). This work was also supported by Health Data Research UK, which is funded by the UK Medical Research Council, the Engineering and Physical Sciences Research Council, the Economic and Social Research Council, the Department of Health and Social Care (England), the Chief Scientist Office of the Scottish Government Health and Social Care Directorates, the Health and Social Care Research and Development Division (Welsh Government), the Public Health Agency (Northern Ireland), the British Heart Foundation and Wellcome. Some analyses were performed using resources provided by the Cambridge Service for Data Driven Discovery (CSD3) operated by the University of Cambridge Research Computing Service (https://www.csd3.cam.ac.uk), provided by Dell EMC and Intel using tier 2 funding from the Engineering and Physical Sciences Research Council (capital grant EP/P020259/1), and DiRAC funding from the Science and Technology Facilities Council (https://www.dirac.ac.uk).

